# Quantitative Ultrasound Biomarkers of Testicular Spermatogenic Function

**DOI:** 10.64898/2026.02.16.26346440

**Authors:** Taylor P. Kohn, Peyton J. Coady, Amelia G. Oppenheimer, Arnaav Walia, Beatriz S. Hernadez, Jaden R Kohn, Niki Parikh, Mahdi Bazzi, Blair T. Stocks, Mohit Khera, Larry I. Lipshultz

## Abstract

**Introduction:** Non-obstructive azoospermia (NOA) represents the most severe form of male infertility. Current clinical tools have limited ability to predict sperm production or guide surgical sperm retrieval. Conventional B-mode ultrasound provides qualitative grayscale images and cannot characterize testicular microstructure relevant to spermatogenesis. Quantitative ultrasound (QUS) provides objective parameters from raw radiofrequency data, which quantitatively measure tissue heterogeneity. We hypothesize that men with spermatogenesis will have different QUS features compared to men without spermatogenesis (measured by total motile count, TMC, on semen analysis), with the goal of identifying imaging biomarkers for prognosis and intraoperative guidance.

**Methods:** We prospectively analyzed men presenting for infertility evaluation who underwent high-frequency ultrasound imaging and semen analysis. Imaging was performed using a 36-MHz transducer with fixed acquisition parameters. Ninety-two QUS features were extracted from manually annotated testicular regions of interest, including Nakagami distribution parameters (m, ω, k), envelope statistics, and texture features. Univariate associations between each QUS feature and TMC were assessed using Spearman correlation with Bonferroni correction. Top-performing features were evaluated using logistic regression and receiver operating characteristic (ROC) analysis to discriminate sperm presence or absence (TMC>0 vs TMC=0).

**Results:** Thirty-seven men (18 azoospermic, 19 with sperm present in the ejaculate) contributed 135 regions of interest. Seventeen of 92 QUS features significantly correlated with TMC after correction. The coefficient of variation of the Nakagami k-factor within the superficial testicular parenchyma (K_Zone1_Cv) demonstrated the strongest correlation (ρ=0.51, corrected p<0.001), suggesting that greater spatial heterogeneity in the superficial parenchyma was associated with higher sperm counts. K_Zone1_Cv discriminated sperm presence with an AUC of 0.77 (95% CI 0.60–0.92), sensitivity 73.7%, and specificity 83.3%. QUS features with the highest univariate association were highly intercorrelated, suggesting a shared biological signal.

**Conclusion:** Quantitative ultrasound–derived measures of testicular microstructure heterogeneity correlate with sperm production and demonstrate moderate discrimination of sperm presence. These findings suggest QUS may provide a non-invasive imaging biomarker of spermatogenesis. Study findings warrant further assessment and validation in male infertility for sperm retrieval prognosis and the potential for intra-operative surgical guidance.

## INTRODUCTION

Non-obstructive azoospermia (NOA) affects approximately 1% of men and represents the most severe form of male infertility.^1^ Successful sperm retrieval during microsurgical testicular sperm extraction (mTESE) ranges from 40–60% depending on the underlying etiology of NOA.^2, 3^ Current methods of clinical assessment – including hormone levels, testicular volume, and conventional testicular ultrasound – provide limited ability to predict which patients will have successful sperm retrieval or to identify which testicular regions are most likely to harbor sperm.^4^ This diagnostic uncertainty leaves patients without reliable prognostic information before mTESE and leaves surgeons without intraoperative guidance, resulting in extensive tissue sampling that may compromise future testicular function.

Conventional ultrasound provides only qualitative grayscale images (B-mode) that reflects the density of anatomical features but cannot characterize the underlying tissue microstructure at a scale relevant to spermatogenesis.^5^ In contrast, quantitative ultrasound (QUS) provides objective parameters from the raw radiofrequency data; these parameters quantitatively measure the properties of the tissue and its acoustic scattering, which allows for a quantifiable interpretation of the size, concentration, organization, and spatial distribution of various tissue components.^6^ QUS parameters have been shown to be clinically useful to characterize tissue microstructure in liver, breast, and other organs, but has been largely unexplored in testicular tissue.^7–9^ In theory, QUS holds significant potential to identify areas of spermatogenesis. The cellular contents within the seminiferous tubules are the fundamental tissue microstructures that are responsible for acoustic scattering in the testis.^10^ We hypothesize that men with active spermatogenesis will have different QUS features compared to men without spermatogenesis. Thus, in this study, we investigated whether various QUS features (Nakagami-based parameters) measured in multiple testicular regions are associated with total motile sperm count (specifically, men with sperm production from those with azoospermia), with the goal of identifying imaging biomarkers for prognosis and intraoperative guidance.

## METHODS

### Study Design

We analyzed men presenting to an academic male fertility clinic between April 2025 to January 2026 under IRB approval. The study cohort included men undergoing evaluation for male infertility who had both high-frequency quantitative ultrasound (HFUS) imaging and semen analysis performed. The primary analysis was an exploration of various QUS features that assess the variation in ultrasound scattering properties in a specific tissue region (measures of local tissue heterogeneity), testing their association with total motile sperm count (TMC) on semen analysis. Laboratory personnel performing semen analysis were unaware of ultrasound results.

Men were excluded if they had a prior testicular sperm extraction procedure (TESE or mTESE), as scar tissue could affect QUS radiofrequency scattering and misclassify tissue heterogeneity.^11^ Men with obstructive azoospermia were also excluded (defined as azoospermia with normal serum FSH ≤7.6 IU/L and normal testicular volume, ≥4.6 cm in length) to ensure the cohort represented men with impaired spermatogenesis, rather than ductal obstruction.^12^

### Ultrasound Acquisition

HFUS imaging was performed by trained operators using a VevoMD high-frequency ultrasound (FUJIFILM VisualSonics, Toronto, Canada) with a 36-MHz linear array transducer with fixed acquisition parameters across all subjects. Raw ultrasound data were acquired in the in-phase and quadrature (IQ) format, allowing direct access to the underlying backscattered signal data rather than relying on vendor-processed images. Imaging depth (~14 mm), lateral field of view (~15.36 mm), gain, and system presets were held constant to minimize technical variability between subjects.

### Ultrasound Radiofrequency Processing

Ultrasound envelope data was reconstructed from the raw IQ (in-phase and quadrature) data using standard magnitude detection. To reduce variability related to coupling, attenuation, and gain, the envelope signal within each ROI was normalized by its median value. This approach is supported by prior work demonstrating that ratio-based normalization preserves relative spatial variation while minimizing the effects of attenuation and system-dependent differences between subjects.^13, 14^

### Quantitative Ultrasound Feature Extraction

QUS analyzes raw radiofrequency echoes to provide objective data about tissue microstructure, unlike conventional B-mode imaging which is qualitative and operator-dependent.^6^ QUS features can be categorized into several families. The ultrasound envelope measures the magnitude of the reflected radiofrequency signal at each point in tissue. QUS statistical features describe the distribution of those amplitudes – including the mean, standard deviation, coefficient of variation, skewness, and kurtosis – differences in the distribution of scattering reflects differences in tissue microstructure.^15^ We extracted 92 QUS features across multiple categories: Nakagami distribution parameters (m-parameter, ω-parameter, k-factor), envelope statistics for each of the Nakagami distribution parameters (mean, coefficient of variation, kurtosis), as well as texture features (gray-level co-occurrence matrix metrics).

### Region of Interest

Regions of interest (ROIs) were manually annotated by a single investigator blinded to semen analysis outcomes. The ROI was drawn to distinguish the testicle from other tissues, such as skin or epididymis, to ensure appropriate regions for analysis. One ROIs was generated per ultrasound image with multiple ultrasound images analyzed per patient. Ultrasound images with excessive acoustic shadowing on B-mode were excluded from analysis. ROIs were defined using the image processing application QuPath (v0.6.0) and exported for quantitative analysis.

On feature analysis, the ROI was divided into three equal depth zones, each comprising one-third of the total ROI depth, as the near-field provides optimal signal-to-noise ratio and minimizes artifacts from depth-dependent acoustic attenuation.^16^

### Statistical Analysis

A comprehensive analysis of 92 QUS features was performed to identify patient-level features associated with the volume of TMC, measured as a continuous variable. Spearman rank correlation coefficients were calculated between patient-level QUS features and TMC. To account for multiple comparisons, Bonferroni correction was applied for 92 comparisons (α = 5.4 × 10^−4^). Complete-case analysis was performed.

After identifying univariate correlation between TMC and each QUS feature, the top 5 QUS features with significant correlation coefficients after Bonferroni correction were then tested to further determine the discriminatory ability of specific QUS features to identify the presence or absence of sperm (rather than quantity of sperm). We performed logistic regression where the outcome of sperm presence was categorized as a binary variable (TMC > 0 versus TMC = 0). Performance metrics included area under the receiver operating characteristic curve (AUC) with bootstrap resampling (2,000 iterations) used to estimate confidence intervals for AUC. For the predictor with the highest AUC, we calculated the sensitivity (probability of correctly identifying men with sperm in the ejaculate) and specificity (probability of correctly identifying men with azoospermia) of that QUS feature.

To assess the degree of redundancy among the top-performing QUS features, we constructed a Spearman correlation matrix. Correlation coefficients were interpreted as weak (|ρ| < 0.4), moderate (0.4 ≤ |ρ| < 0.7), strong (0.7 ≤ |ρ| < 0.9), or very strong (|ρ| ≥ 0.9). All QUS image processing and statistical analyses were performed in Python.

## RESULTS

### Study Cohort and Clinical Characteristics

A total of 37 men were included, contributing 135 regions of interest across multiple testicular sites (median 5 ROIs per patient). Of these, 18 men (49%) had azoospermia (TMC = 0) and 19 men (51%) had sperm present on semen analysis (TMC > 0) (Supplemental Figure 1). Among men with sperm present, TMC ranged from 0.03 to 30.42 million (median 7.7 million). Endocrine and semen characteristics are summarized in Table 1.

**Table 1.** Clinical Characteristics of Included Men.

| **Variable** | **Azoospermia** | **Sperm In Ejaculate** |
| --- | --- | --- |
| Number of Men | 18 | 19 |
| FSH (mIU/mL) | 18.9 (13.8-26.2) | 4.0 (2.5-7.7) |
| LH (mIU/mL) | 9.5 (7.5-14.1) | 3.7 (2.7-5.1) |
| Testosterone (ng/dL) | 295.5 (225.8-318.2) | 352.0 (240.8-422.8) |
| Volume (mL) | 2.0 (0.9-4.9) | 2.5 (2.0-3.5) |
| Concentration (M/mL) | 0.0 (0.0-0.0) | 4.6 (0.5-19.2) |
| Motility (%) | 0.0 (0.0-0.0) | 40.0 (0.0-50.0) |
| TMC (million motile sperm) | 0.0 (0.0-0.0) | 6.6 (1.5-16.9) |

Baseline endocrine, and semen parameters of men undergoing quantitative ultrasound (QUS) imaging and semen analysis. Continuous variables are presented as median (interquartile range). The cohort includes men with azoospermia (TMC = 0) and men with sperm present in the ejaculate (TMC > 0). Hormonal parameters include follicle-stimulating hormone (FSH), luteinizing hormone (LH), total testosterone, and other available reproductive markers.

### Comprehensive QUS Feature Analysis

Of 92 QUS features analyzed, 17 features (18%) demonstrated significant correlation with TMC after Bonferroni correction (Table 2). K_Zone1_Cv (the coefficient of variation of the Nakagami k-factor for the superficial parenchyma) demonstrated the strongest correlation with TMC among all features analyzed (Spearman ρ = +0.51, corrected p = 5.71 × 10^−8^) (Figure 1A). The Nakagami k-factor describes the magnitude of the acoustic scattering. The coefficient of variation (CV) represents the spread of values compared with the average (standard deviation divided by mean). A low CV indicates the data is clustered near the average, whereas high CV indicates the data are spread out compared to the average. Thus, the CV of the k-factor represents spatial heterogeneity. If the tissue is very heterogeneous, then the acoustic scattering varies significantly within the ROI (high CV), whereas if the tissue is very homogeneous, then the acoustic scattering is more uniform within the ROI (low CV). Thus, positive correlation between K_Zone1_Cv and TMC suggests that greater spatial heterogeneity in the superficial testicle parenchyma is associated with higher sperm counts.

**Table 2.** The 25 Quantitative Ultrasound Features Associated with Total Motile Counts.

| Rank | Feature | Spearman Coefficient | P Value (Uncorrected) | P Value (Bonferroni Correction) |
| --- | --- | --- | --- | --- |
| 1 | k_zone1_cv | 0.51 | 6.20E-10 | 5.71E-08 |
| 2 | m_zone1_median | -0.46 | 1.64E-08 | 1.51E-06 |
| 3 | m_cv | 0.44 | 6.66E-08 | 6.13E-06 |
| 4 | k_zone1_median | -0.43 | 1.92E-07 | 1.77E-05 |
| 5 | k_zone1_iqr | -0.42 | 3.14E-07 | 2.89E-05 |
| 6 | m_zone1_mean | -0.42 | 4.38E-07 | 4.03E-05 |
| 7 | k_zone1_mean | -0.41 | 1.01E-06 | 9.27E-05 |
| 8 | m_zone2_cv | 0.39 | 2.40E-06 | 2.21E-04 |
| 9 | m_p10 | -0.39 | 2.78E-06 | 2.55E-04 |
| 10 | m_zone1_cv | 0.39 | 3.65E-06 | 3.36E-04 |
| 11 | k_zone2_cv | 0.37 | 1.16E-05 | 1.07E-03 |
| 12 | k_across_zones_cv | 0.36 | 1.88E-05 | 1.73E-03 |
| 13 | m_median | -0.36 | 2.33E-05 | 2.14E-03 |
| 14 | m_zone3_cv | 0.35 | 2.55E-05 | 2.35E-03 |
| 15 | omega_median | 0.33 | 7.91E-05 | 7.28E-03 |
| 16 | k_cv | 0.33 | 9.48E-05 | 8.72E-03 |
| 17 | m_zone2_median | -0.33 | 1.03E-04 | 9.48E-03 |
| 18 | m_mean | -0.29 | 5.74E-04 | 5.28E-02 |
| 19 | m_across_zones_cv | 0.28 | 8.88E-04 | 8.17E-02 |
| 20 | m_zone2_mean | -0.28 | 1.01E-03 | 9.25E-02 |
| 21 | omega_zone1_median | 0.27 | 1.77E-03 | 1.63E-01 |
| 22 | omega_iqr | 0.26 | 2.07E-03 | 1.91E-01 |
| 23 | env_p10 | -0.26 | 2.12E-03 | 1.95E-01 |
| 24 | k_zone3_cv | 0.26 | 2.67E-03 | 2.45E-01 |
| 25 | k_zone2_median | -0.25 | 2.96E-03 | 2.72E-01 |

Ranked list of the 25 QUS features (of 92 analyzed) demonstrating the strongest Spearman correlation coefficients with total motile sperm count (TMC). Features are ordered by absolute magnitude of correlation ( $|\rho|$ ). Bonferroni-corrected p-values are shown. Feature families include Nakagami distribution parameters (m, $\omega$ , k), envelope statistical measures (mean, coefficient of variation, kurtosis), and texture metrics derived from gray-level co-occurrence matrices.

**Table 3.**
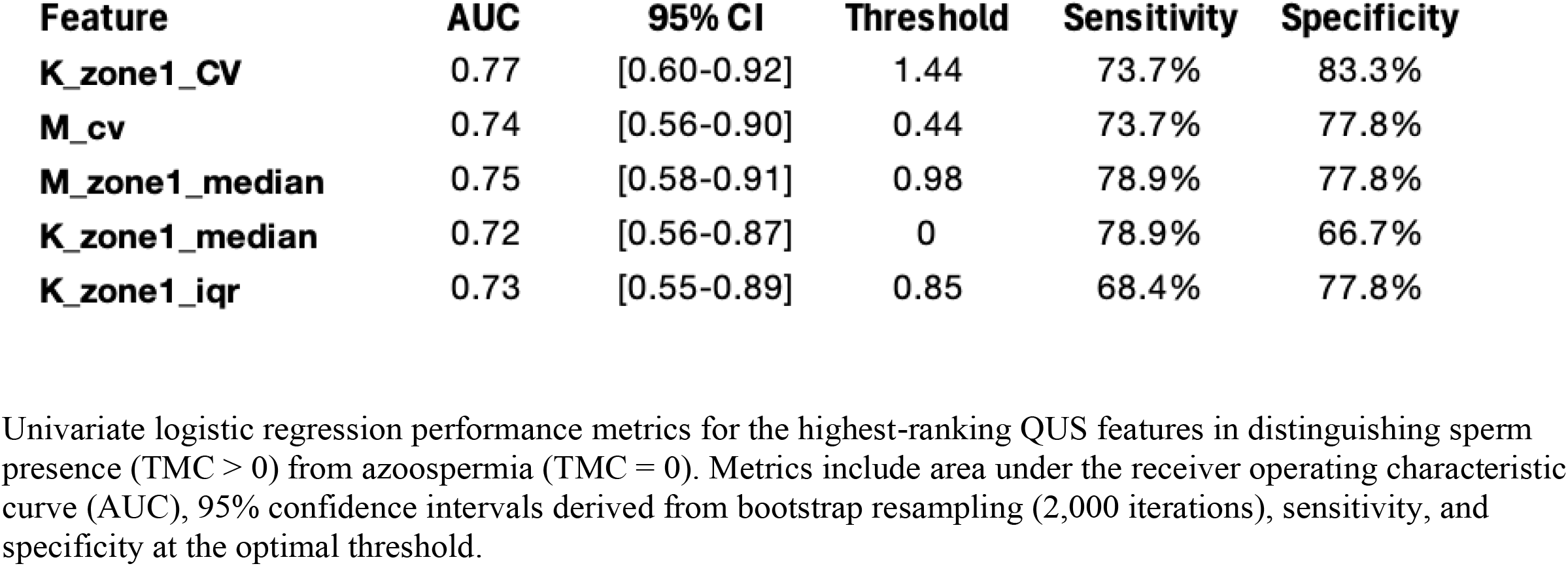
Univariate logistic regression performance metrics.

**Figure 1.**
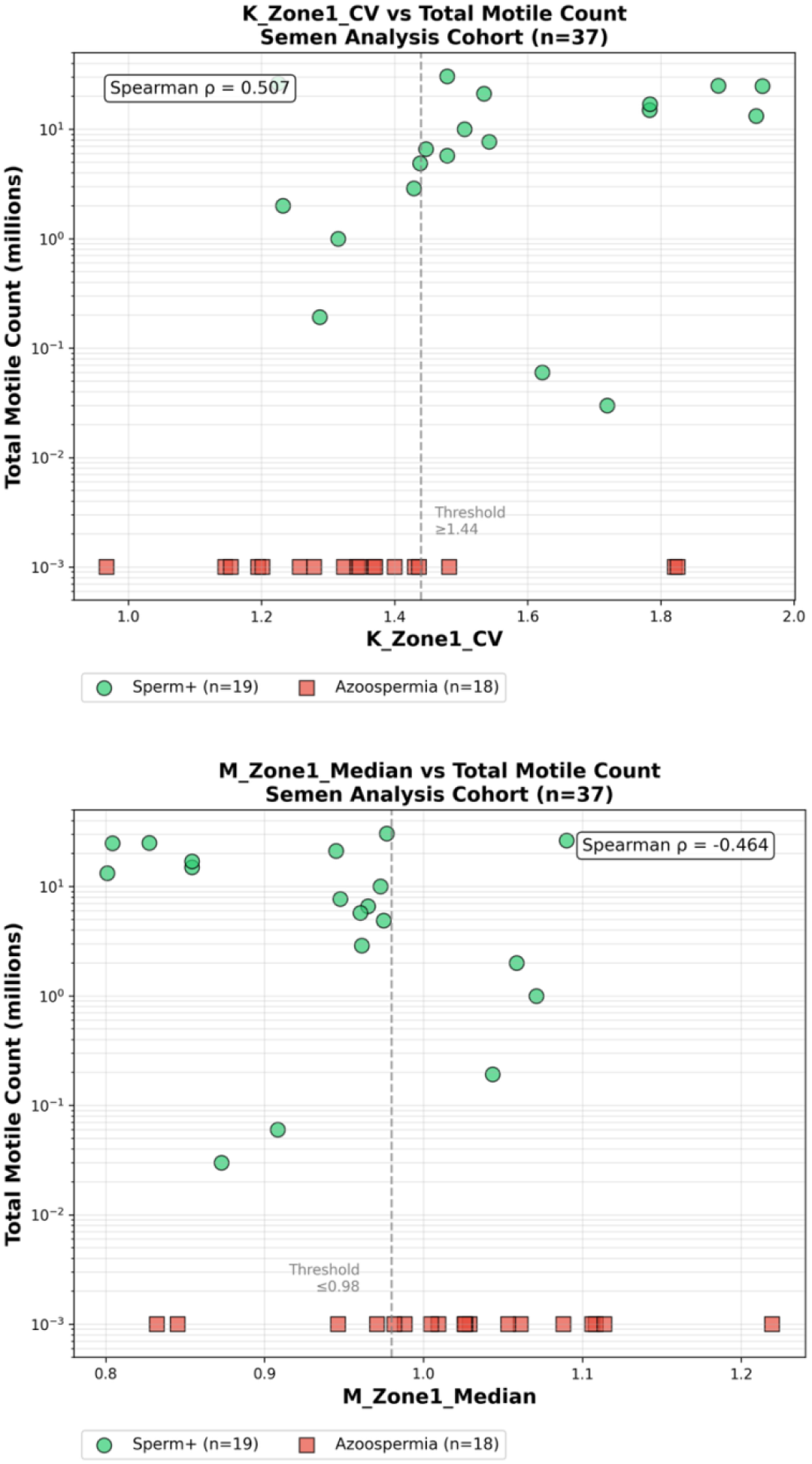
Scatter Plot for K_Zone1_Cv and M_Zone1_Median **(A)** Scatter plot demonstrating the positive correlation between K_Zone1_Cv (coefficient of variation of the superficial Nakagami k-factor) and total motile sperm count (TMC). Greater spatial heterogeneity in the superficial testicular parenchyma is associated with higher sperm counts. Bonferroni-corrected p-value = 5.71 × 10^−8^. **(B)** Scatter plot demonstrating the negative correlation between m_Zone1_Median and TMC. Lower median Nakagami m-values (greater heterogeneity) are associated with higher sperm counts. Bonferroni-corrected p-value = 1.51 × 10^−6^.

The second strongest correlation was m_zone1_median (ρ = −0.462, corrected p = 1.51 × 10^−6^). An m-value <1 suggests highly heterogeneous tissue, whereas an m-value >1 suggests uniform tissue. A negative correlation between TMC and the median of the m-value within the superficial testicle parenchyma suggests that more uniform tissue is associated with lower sperm counts (Figure 1B).

### Univariate Discrimination of Sperm Presence

K_Zone1_Cv discriminated men with sperm present from men with azoospermia with an AUC of 0.77 (95% CI: 0.60 - 0.92) in univariate analysis, with a sensitivity of 73.7% and specificity of 83.3% (Table . Men with sperm present had significantly higher K_Zone1_Cv values compared to men with azoospermia (1.56 ± 0.22 vs. 1.36 ± 0.21; Mann-Whitney U p = 0.005, Figure 2). M_CV discriminated men with sperm present from men with azoospermia with an AUC of 0.74 (95% CI: 0.56 - 0.90).

**Figure 2.**
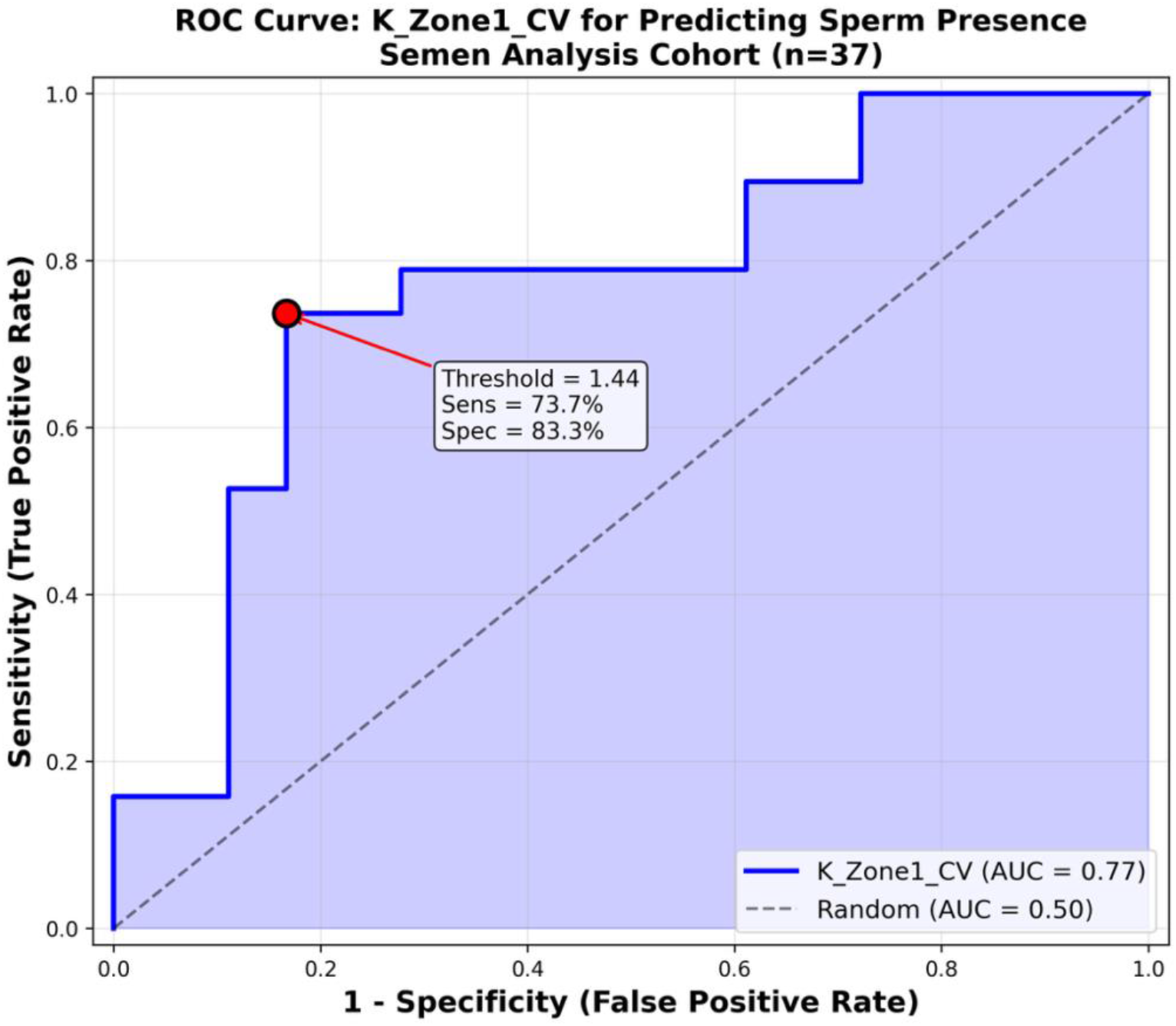
ROC Curves for Classification of Sperm Presence in Ejaculate vs Azoospermia Receiver operating characteristic (ROC) curves demonstrating the ability of top QUS features to classify sperm presence versus azoospermia. K_Zone1_Cv demonstrated the highest discriminatory performance (AUC 0.77, 95% CI 0.60–0.92). Curves illustrate sensitivity–specificity tradeoffs across threshold values.

### Spearman Coefficient Feature Correlation

The five top-ranked QUS features demonstrated high intercorrelation, with Spearman coefficients ranging from |ρ| = 0.58 to 0.98 (Table X). The strongest correlation was observed between K_Zone1_Cv and M_Zone1_Median (ρ = −0.98), indicating these parameters capture nearly identical biological information despite measuring different acoustic properties. Similarly strong correlations were found between many of these top parameters (Supplemental Table 1). Given this high degree of multicollinearity, K_Zone1_Cv was selected as the single representative biomarker for subsequent analyses to avoid redundancy and simplify clinical interpretation.

## DISCUSSION

This is the first study to explore the use of quantitative ultrasound in male infertility. Quantitative ultrasound offers numerous potential benefits as a noninvasive method with the potential for real-time application of complex radiofrequency data to characterize testicular microstructure beyond what conventional B-mode imaging can detect; QUS parameters use raw radiofrequency data and transform this into meaningful metrics that describe tissue organization at a scale relevant to spermatogenesis.^17^ Currently, active spermatogenesis is indistinguishable using conventional B-mode ultrasound.^18^ This initial foray into QUS within male infertility examined a simple analysis, exploring the correlation between the presence of motile sperm in the ejaculate with a wide net of QUS parameters. A few parameters were able to be identified that characterize tissue heterogeneity and were associated with sperm presence on semen analysis. This phenomenon has biological plausibility, as areas of active spermatogenesis on mTESE have histologic findings that suggest heterogeneity among tubules in various stages of spermatogenesis, whereas men with azoospermia demonstrate sclerotic tubules, which are more uniform in their microarchitecture.^19, 20^ While this analysis represents extremely early work in the field of QUS application to male infertility, it is reasonable to hypothesize that (after further development of the mathematical models) a future exists in which real-time QUS analysis can be applied to guide surgical sperm retrieval – using pre-operative QUS to provide an accurate prognosis for the patient and then using QUS intra-operatively to target regions most likely to harbor sperm, which could reduce operative time, minimize tissue disruption, and improve retrieval success rates.

Quantitative ultrasound analyses have been used clinically in several areas of medicine. Quantitative ultrasound can be used for osteoporosis assessment and fracture risk prediction with efficiency comparable to X-ray densitometry as US parameters reflect both bone mineral density and trabecular microarchitecture. QUS techniques are increasingly used for chronic liver disease evaluation as these analyses can detect fat accumulation, inflammation, and fibrosis through changes in acoustic properties.^21^ Finally, QUS has demonstrated high accuracy in distinguishing benign from malignant breast lesions, with multiple studies reporting AUC values exceeding 0.90 as QUS parameters reflect cellular organization, nuclear characteristics, stromal composition, and vascular properties.^22, 23^ These examples demonstrate that QUS can be used to detect differences in tissue microstructures that exist in histopathologic states.

We hypothesize that QUS can also be helpful in distinguishing between healthy, active spermatogenesis and pathologic, sclerotic tubules without spermatogenesis. Healthy sperm production creates a mix of tubules in different stages — some full of sperm, some earlier in development, some resting — so the tissue looks varied under quantitative ultrasound. K_Zone1_Cv measures that variability: high values indicate heterogeneity with a complex patchwork of activity, while low values suggest homogeneity – uniform appearing tubules.

While this study is a novel application of quantitative ultrasound in the setting of male infertility, this study has several limitations that should be considered when interpreting the findings. First, this was an exploratory analysis to determine if features of QUS correlated with variations in sperm production that may have some biological rationale. The sample size was small (n=37), and the cohort was dichotomized into azoospermic (n=18) and sperm-positive (n=19) groups for classification analysis so limited clinical utility can be derived. As an exploratory pilot study; no formal sample size calculation was performed. While this enabled evaluation of diagnostic performance, the limited sample precludes robust multivariable modeling and may not capture the full spectrum of testicular pathology. Additionally, among men with a diagnosis of non-obstructive azoospermia, none had undergone sperm retrieval procedures at the time of analysis, so the model could be detecting features associated with spermatogenesis that are not making it into the ejaculate. Future studies comparing NOA men with sperm retrieval results are ongoing.

Further, the number of ROIs varied across patients, introducing potential sampling heterogeneity. Although patient-level analyses used median values across ROIs to mitigate this, unequal sampling may influence feature stability, particularly for patients with fewer interrogated sites. Standardization of ROI acquisition protocols will be important for clinical translation. Next, the QUS features were derived from a single cross-sectional imaging session. Intra-patient reproducibility - including test-retest reliability, inter-operator variability, and sensitivity to probe positioning and pressure - was not formally assessed in this study. Establishing measurement reproducibility is a prerequisite for clinical adoption. Finally, the probability estimates used for clinical interpretation were derived from quadrant analysis of the same cohort and should be considered preliminary. External validation in independent populations - including men with normal fertility, prior testicular surgery, and varied etiologies - is necessary before these thresholds can be applied in clinical decision-making.

In this exploratory study, quantitative ultrasound analysis identified tissue heterogeneity patterns that differentiated men with and without sperm. These early findings suggest that QUS may capture meaningful differences in testicular microstructure, but the small sample sizes and lack of histological confirmation limit the conclusions that can be drawn. Prospective validation in larger populations is essential before any clinical application can be considered.

## Supporting information

STARD Checklist

## Data Availability

All data produced in the present study are available upon reasonable request to the authors

---

**Supplemental Figure 1:**
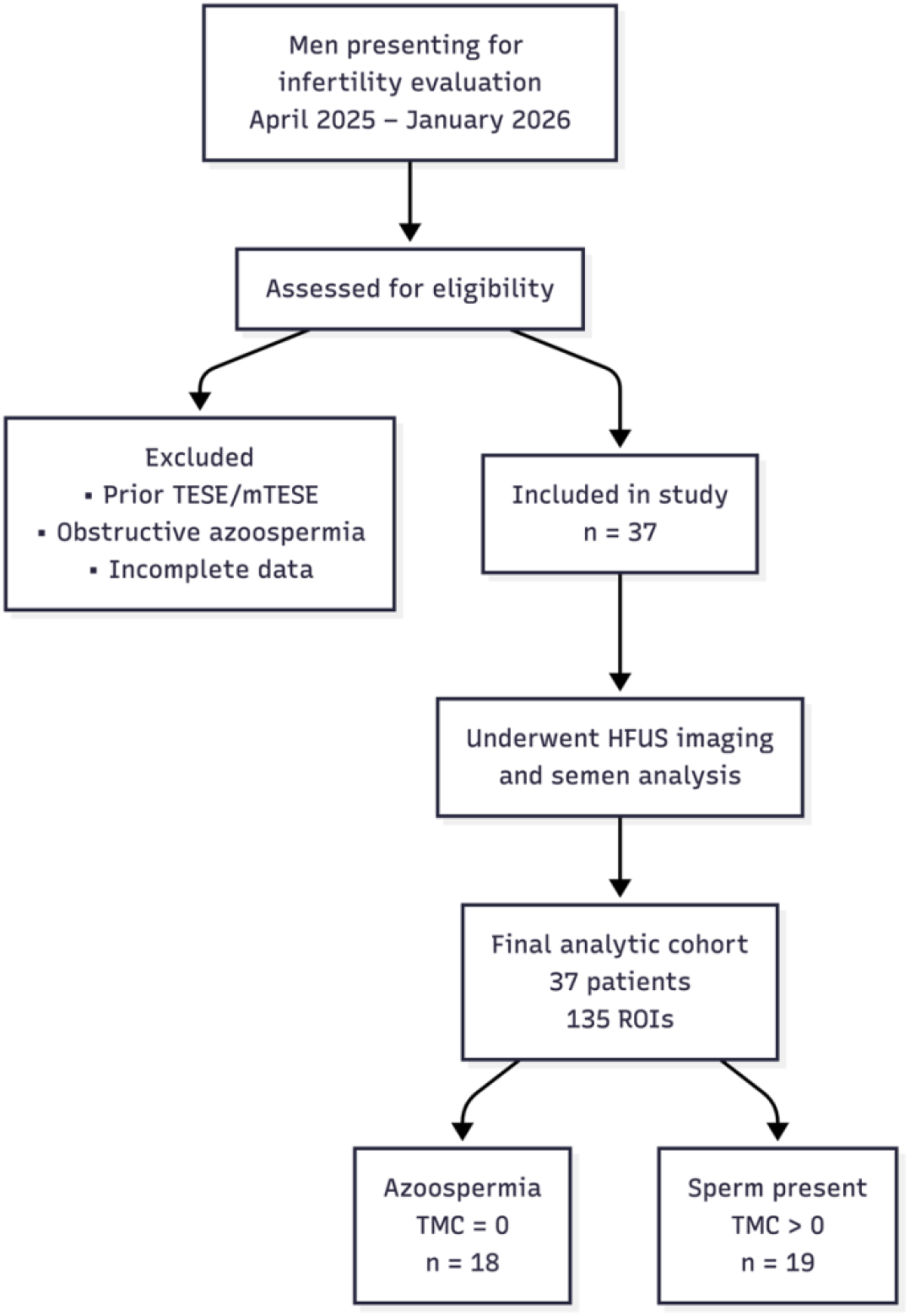
STARD Flow Diagram of Study Participants Flow diagram of participant inclusion and diagnostic classification in this prospective study evaluating quantitative ultrasound (QUS) biomarkers of spermatogenesis. Men presenting for infertility evaluation between April 2025 and January 2026 underwent high-frequency quantitative ultrasound (HFUS) imaging and semen analysis during the same clinical assessment. Thirty-seven men met the inclusion criteria and comprised the final analytic cohort, contributing 135 regions of interest (ROIs). Participants were categorized according to total motile sperm count (TMC) as azoospermic (TMC = 0; n = 18) or sperm present (TMC > 0; n = 19). Abbreviations: HFUS, high-frequency ultrasound; ROI, region of interest; TMC, total motile sperm count.

**Supplemental Table 1.** Semen Quantitative Ultrasound Feature Correlation Matrix.

|  | **k_zone1_cv** | **m_zone1_median** | **k_zone1_median** | **m_cv** | **k_zone1_iqr** |
| --- | --- | --- | --- | --- | --- |
| **k_zone1_cv** | 1 | -0.98 | -0.9 | 0.74 | -0.94 |
| **m_zone1_median** | -0.98 | 1 | 0.9 | -0.68 | 0.97 |
| **k_zone1_median** | -0.9 | 0.9 | 1 | -0.64 | 0.88 |
| **m_cv** | 0.74 | -0.68 | -0.64 | 1 | -0.58 |
| **k_zone1_iqr** | -0.94 | 0.97 | 0.88 | -0.58 | 1 |

Spearman correlation matrix demonstrating intercorrelation among the five highest-ranking QUS features associated with TMC. Correlation strength is categorized as weak ( $|\rho| < 0.4$ ), moderate (0.4–0.7), strong (0.7–0.9), or very strong ( $>0.9$ ). High multicollinearity among top parameters suggests shared biological signal reflecting tissue heterogeneity.

## REFERENCES

1. Cerván-Martín M, Castilla JA, Palomino-Morales RJ, Carmona FD. Genetic Landscape of Nonobstructive Azoospermia and New Perspectives for the Clinic. J Clin Med. 2020;9.

2. Schlegel PN. Testicular sperm extraction: microdissection improves sperm yield with minimal tissue excision. Hum Reprod. 1999;14:131–135.

3. Corona G, Minhas S, Giwercman A, et al. Sperm recovery and ICSI outcomes in men with non-obstructive azoospermia: a systematic review and meta-analysis. Hum Reprod Update. 2019;25:733–757.

4. Brannigan RE, Hermanson L, Kaczmarek J, Kim SK, Kirkby E, Tanrikut C. Updates to Male Infertility: AUA/ASRM Guideline (2024). J Urol. 2024;212:789–799.

5. Popp RL, Macovski A. Ultrasonic diagnostic instruments. Science. 1980;210:268–273.

6. Oelze ML, Mamou J. Review of Quantitative Ultrasound: Envelope Statistics and Backscatter Coefficient Imaging and Contributions to Diagnostic Ultrasound. IEEE Trans Ultrason Ferroelectr Freq Control. 2016;63:336–351.

7. Zhang LX, Dioguardi Burgio M, Vilgrain V, et al. Quantitative Ultrasound and Ultrasound-Based Elastography for Chronic Liver Disease: Practical Guidance, From the AJR Special Series on Quantitative Imaging. AJR Am J Roentgenol. 2025;225:e2431709.

8. Sadeghi-Naini A, Suraweera H, Tran WT, et al. Breast-Lesion Characterization using Textural Features of Quantitative Ultrasound Parametric Maps. Sci Rep. 2017;7:13638.

9. Ashir A, Jerban S, Barrère V, et al. Skeletal Muscle Assessment Using Quantitative Ultrasound: A Narrative Review. Sensors (Basel). 2023;23.

10. Giffin JL, Bartlewski PM, Hahnel AC. Correlations among ultrasonographic and microscopic characteristics of prepubescent ram lamb testes. Exp Biol Med (Maywood). 2014;239:1606–1618.

11. Luchies AC, Ghoshal G, O’Brien WD, Jr., Oelze ML. Quantitative ultrasonic characterization of diffuse scatterers in the presence of structures that produce coherent echoes. IEEE Trans Ultrason Ferroelectr Freq Control. 2012;59:893–904.

12. Schoor RA, Elhanbly S, Niederberger CS, Ross LS. The role of testicular biopsy in the modern management of male infertility. J Urol. 2002;167:197–200.

13. Destrempes F, Franceschini E, Yu FT, Cloutier G. Unifying Concepts of Statistical and Spectral Quantitative Ultrasound Techniques. IEEE Trans Med Imaging. 2016;35:488–500.

14. Mori S, Montobbio N, Sormani MP, et al. Echocardiographic Tissue Characterization Using Radiomics in Patients With Transthyretin-Related Cardiac Amyloidosis. JACC Adv. 2025;4:101755.

15. Tehrani AKZ, Cloutier G, Tang A, Rosado-Mendez IM, Rivaz H. Homodyned K-Distribution Parameter Estimation in Quantitative Ultrasound: Autoencoder and Bayesian Neural Network Approaches. IEEE Trans Ultrason Ferroelectr Freq Control. 2024;71:354–365.

16. Szabo TL. Diagnostic ultrasound imaging : inside out. Second edition ed. Oxford: Elsevier/Academic Press; 2014.

17. Huang WL, Ding L, Yao JH, et al. Testicular quantitative ultrasound: A noninvasive monitoring method for evaluating spermatogenic function in busulfan-induced testicular injury mouse models. Andrologia. 2021;53:e13927.

18. Ohta T, Kojo K, Kurobe M, et al. Feasibility of high-frequency ultrasound for seminiferous tubule assessment and correlation of B-mode imaging with pathological findings in the testis in azoospermia. J Med Ultrason (2001). 2024;51:465–475.

19. Punjani N, Flannigan R, Kang C, Khani F, Schlegel PN. Quantifying Heterogeneity of Testicular Histopathology in Men with Nonobstructive Azoospermia. J Urol. 2021;206:1268–1275.

20. Paniagua R, Nistal M, Amat P, Rodriguez MC, Martin A. Seminiferous tubule involution in elderly men. Biol Reprod. 1987;36:939–947.

21. Hans D, Métrailler A, Gonzalez Rodriguez E, Lamy O, Shevroja E. Quantitative Ultrasound (QUS) in the Management of Osteoporosis and Assessment of Fracture Risk: An Update. Adv Exp Med Biol. 2022;1364:7–34.

22. Wang C, Che Y. A ultrasonic nomogram of quantitative parameters for diagnosing breast cancer. Sci Rep. 2023;13:12340.

23. Osapoetra LO, Chan W, Tran W, Kolios MC, Czarnota GJ. Comparison of methods for texture analysis of QUS parametric images in the characterization of breast lesions. PLoS One. 2020;15:e0244965.

