## Supplementary material for "Quantitative Ultrasound Biomarkers of Testicular Spermatogenic Function": STARD Checklist

### STARD 2015 Checklist

1–2. Title/Abstract: Identified as diagnostic accuracy study; structured abstract with AUC, sensitivity, specificity reported.

3–4. Introduction: Scientific background and hypothesis clearly stated.

5. Study Design: Prospective clinical cohort (April 2025–January 2026).

6–9. Participants: Inclusion/exclusion criteria defined; obstructive azoospermia and prior TESE excluded.

10a. Index Test: High-frequency quantitative ultrasound (36-MHz transducer); raw IQ processing; 92 QUS features extracted.

10b. Reference Standard: Semen analysis measuring total motile sperm count (TMC).

11. Rationale: TMC used as biologic proxy for spermatogenesis.

12a–12b. Cutoffs: Continuous QUS features analyzed; binary outcome defined as TMC > 0 vs TMC = 0.

13a–13b. Blinding: ROI annotation blinded to semen results; laboratory personnel blinded to ultrasound findings.

14–16. Statistical Analysis: Spearman correlation with Bonferroni correction; logistic regression; ROC/AUC with bootstrap CIs; complete-case analysis.

17. Variability Analysis: Superficial vs deeper ROI zones evaluated.

18. Sample Size: Exploratory pilot study; no formal sample size calculation performed.

19–21. Participant Flow & Baseline Data: STARD flow diagram provided; baseline endocrine and semen characteristics reported.

22. Timing: Imaging and semen analysis performed during same clinical evaluation period.

23–24. Diagnostic Accuracy: AUC 0.77 (95% CI 0.60–0.92); sensitivity 73.7%; specificity 83.3%.

25. Adverse Events: None occurred.

26–27. Limitations & Clinical Implications: Small sample; no histologic validation; need for external validation.

28–30. Other Information: IRB approval obtained; no funding; no conflicts of interest.
